# Service user perspectives of community mental health services for people with complex emotional needs: a co-produced qualitative interview study

**DOI:** 10.1101/2021.06.24.21259476

**Authors:** Kylee Trevillion, Ruth Stuart, Josephine Ocloo, Eva Broeckelmann, Stephen Jeffreys, Tamar Jeynes, Dawn Allen, Jessica Russell, Jo Billings, Mike J Crawford, Oliver Dale, Rex Haigh, Paul Moran, Shirley McNicholas, Vicky Nicholls, Una Foye, Alan Simpson, Brynmor Lloyd-Evans, Sonia Johnson, Sian Oram

## Abstract

**Background:** There is consensus that services supporting people with complex emotional needs are part of a mental health care system in which change is needed. To date, service users’ views and co-production exercises have had little impact on the development of treatment and care. This needs to change, and our paper evidences the experiences and perspectives of a diverse range of people on how community services can best address the needs of people with complex emotional needs.

**Methods:** A co-produced qualitative research study. Lived experience researchers led data collection and analysis. Individual interviews were conducted with 30 people across England who had a diverse range of experiences and perspectives of using community services for complex emotional needs. Participants were asked about their experiences of using community services for their mental health, and views on how community services can best address their needs. Thematic analysis was used to analyse the data.

**Results:** Participants reported some experiences of good practice but also of experiences of severely stigmatising treatment, a lack of effective support and service fragmentation. *Relational Practice* was identified as the central overarching theme and describes how community services can best support people with complex emotional needs. This approach involves care delivered in a non-stigmatising, individualised, compassionate and trauma-informed manner. It involves care that is planned collaboratively with service users to ensure their multiple needs are addressed in a flexible, holistic and consistent way which accounts for the long-term and fluctuating nature of their needs.

**Conclusions:** Relational practice approaches have potential to facilitate better community care for people with complex emotional needs. Research and service development are needed to examine how best to implement such approaches across the mental health service system. This work must be co-produced with people with relevant lived experience, their carers and the professionals who support them.

## Background

A recent systematic review examined the worldwide prevalence of any ‘personality disorder’ across 21 countries and estimated that up to 8% of community populations are affected (1). Higher rates are found within community healthcare settings, with around a quarter of people accessing primary care services and half of people accessing outpatient mental health services meeting criteria for a ‘personality disorder’ (2, 3). Evidence indicates that the prevalence of ‘personality disorder’ in the general community is similar among men and women and as common among minority ethnic groups as majority groups (3, 4). Yet, in clinical populations the prevalence of ‘personality disorder’ diagnoses are lower among minority ethnic groups and among men. It is unclear whether these differences reflect a lower prevalence meeting diagnostic criteria or instead lower rates of service use and/or under-detection by services (3, 5).

Our team recognises the considerable stigma attached to the label ‘personality disorder’, and the considerable associated harms identified by both service users and clinicians (6, 7). Many people find it unhelpful and do not identify with it. For this reason, in this paper and in our study materials we used a working term that is also used by some mental health services in the UK - ‘complex emotional needs’ (CEN) - to delineate the group of service users who may have received a ‘personality disorder’ diagnosis and/or have used services for ‘personality disorder’ or CEN, or who appear to have similar needs (e.g. related to repeated self-harm). It is not our intention that complex emotional needs becomes a substitute diagnosis, but rather a description of a broad group of service users and survivors. We advocate co-produced work to develop new ways of describing and assessing their difficulties.

Mental health services are found to marginalise service users with CEN and, compared with other mental health conditions, the provision of timely, well-resourced interventions and good quality care for this service user population appears to lag behind (7–10). Our recent meta-synthesis of the international evidence on service users’ experiences of mental health services identified several areas for which there is a strong consensus on what kind of care is needed (6). These include providing holistic support (i.e. support that addresses service users’ psychological, social, and physical needs), delivered by skilled and compassionate staff who understand the need for a long-term perspective on treatment. Our complementary meta-synthesis of international clinician perspectives highlights that some staff, especially in generic mental health services, hold ambivalent, pessimistic, and stigmatising views about people with CEN and may lack the knowledge and skills to effectively support people (7). Access to specialist services and longer-term treatments are also reported to be impeded by a lack of clear referral pathways and accessible services for people at various stages in recovery journeys (7). Clinicians call for better organisational support, more joint-working practices and clinical supervision to assist them in delivering better care to service users (7).

Given the high levels of need within community services and the considerable variability in service quality, a growing number of international policy guidelines are intended to improve and enhance community care for people with CEN (11, 12). Yet, the data sources used to develop many of these guidelines fail to incorporate the views and perspectives of service users and the family and friends who support them (11, 13). In addition, most research studies on care models for people with CEN focus on psychotherapeutic treatment models, especially those intended to reduce self-harm (14). They ignore the more general principles of how to provide good care and ensure peoples’ needs are met across healthcare service systems. In this co-produced qualitative study, our team of researchers and clinicians, including several with relevant lived experience, aimed to identify best practice in community treatment and support for people with CEN from the perspective of services users. This study contributes to foundations for intervention development and service improvements that are informed by service user perspectives and priorities. The study objectives were:

1. To explore the experiences of adults with CEN in using community services for their mental health in a range of English service settings, including National Health Service (NHS) and voluntary sector services
2. To explore the views of adults with CEN about how community services can best meet their needs

## Methods

We followed CORE-Q reporting guidelines for qualitative research (15).

### Study sample

People were eligible for the study if they were: adults (aged 18 or above) who had received a diagnosis of a ‘personality disorder’ or who self-identified as having difficulties that may result in a ‘personality disorder’ diagnosis or in using CEN services (e.g. recurrent self-harm, other impulsive behaviour); who may have used community services for their mental health; who could undertake an interview in English; who had capacity to consent to the research. We recruited 30 people and aimed to ensure our sample represented a full range of characteristics with respect to age, gender, sexuality, ethnicity, geographical location in England and type of community service use. We constantly reviewed the sample, as a working group, to monitor its representativeness. We adopted an intersectional (i.e. recognizing individuals’ intersecting identities including race, class, gender, sexuality, disability) and culturally competent (i.e. considering cultural identity and context) approach to recruitment to capture a diversity of perspectives.

### Study definition

We defined community services as: (1) publicly-funded primary care services, which provide the first point of contact in the UK healthcare system (e.g. general practitioner services); (2) publicly-funded non-specialist secondary mental health services (e.g. community mental health teams) and specialist community ‘personality disorder’ services, which are accessed via referral from primary care and generic mental health services; (3) non-profit community organisations and networks whose remit involves face-to-face work with people with CEN. Community forensic mental health services fell outside our study definition of community services.

### Recruitment

Participants were recruited from voluntary sector organisations for people with mental health problems (e.g. National Survivor User Network), including those for Black communities and Lesbian, Gay, Bisexual, Transgender, Queer/Questioning people (LGBTQ+) and relevant online social media networks (e.g. the Mental Health Policy Research Unit, other personal and institutional Twitter accounts, the Mental Elf Twitter account, Facebook accounts on mental health). We used advertisements to recruit people and developed these to promote engagement of under-represented groups (e.g. including images of Black men and women and rainbow symbols for LGBTQ+ people). People interested in the study could either contact the research team directly, using the contact information on the advert, or ask that their information be passed on to the research team via the network managers/coordinators. One of the study researchers (KT, JO) had an initial conversation with people who were interested in the study, to establish whether they met eligibility criteria and to ensure diversity was achieved in the sample of people who were interviewed. Eligible participants were sent copies of the Participant Information Sheet at least 24 hours before the interview. Participants were offered the option of being interviewed alone or with someone else present with them at the interview (e.g. a close friend or family member).

### Data Collection

Recruitment occurred between July 2019-October 2020 (N.B. the study was suspended between March and July 2020 in response to the COVID-19 pandemic). Before COVID-19, participants were given the option of being interviewed by a researcher either face-to-face, by Skype or telephone. Face-to-face research interviews were conducted within university settings. During COVID-19, with social distancing requirements in place, all interviews were conducted remotely using MS Teams or Zoom video-conferencing software; interviews were conducted by a researcher and a facilitator who was responsible for the interview recording. Informed consent was obtained from all participants prior to the interview, either in written form or verbally recorded. All interviews were recorded (either on an encrypted digital recorder or laptop) and were transcribed verbatim. The length of interviews varied from 30-100 minutes.

### Research Team

This study was co-produced from inception, design and delivery by members of the Mental Health Policy Research Unit (KT, RS, JO, UF, SJo, AS, BLE, VN, SO) and a group of six experts by experience (SJe, EB, TJ, DA, JR and Gabriella Clarke) and nine experts by occupation (OD, PM, MC, RH, SMc, JB, Alison Bearn, Brian Solts, Penny Bennett). Work included the co-production of the study protocol, interview topic guide and analyses. Five of the six experts by experience (SJe, EB, TJ, DA and JR) conducted most of the research interviews (n=24) and four of them (EB, SJe, DA, JR) coded most of the interview transcripts (n=20). Six interviews were conducted by a health researcher (KT) and the remaining transcripts were coded by five health researchers (Nafiso Ahmed, Norha Vera San Juan, RS, KT, UF). Two health researchers adapted the study documentation and interview procedures following the pandemic (JO, RS). Of the paper authors, seven are male, and two have Black or mixed heritage backgrounds.

### Analysis

Thematic analysis (Braun and Clarke 2013) was used, and the data managed in NVivo Pro V12 (16). Six key steps were undertaken. The first step, familiarisation with the data, involved detailed readings of the interview transcripts by researchers, who made initial reflections/notes about the narratives. The second step, generation of initial codes, involved line-by-line open coding of each transcript. The team first did this as a group, using one transcript to establish an initial coding frame. The remaining transcripts were then shared out among researchers to complete the third, fourth and fifth steps of the analyses, involving searching for, reviewing, and defining the arising themes; the coding frame was revised and updated accordingly. The sixth step involved categorisation of data into a final set of themes which were conceptualised in a thematic map.

## Results

### Sample

We recruited 30 adults with diverse characteristics of age, ethnicity, geographical location and use of community services. Although we aimed to recruit some people who had not received a ‘personality disorder’ diagnosis, all participants had received a diagnosis at some point in their life. Some diversity was achieved with gender and sexuality (See Table 1).

**Table 1.** Study Sample Characteristics.

| Primary sampling criteria | Category | N |
| --- | --- | --- |
| Current diagnoses* | Personality disorder | 29 |
|  | Complex post-traumatic stress disorder | 1 |
| Service use | Specialist | 16 |
|  | Non-specialist | 10 |
|  | Unclear if specialist or non-specialist | 4 |
| Age | 18-24 | 3 |
|  | 25-34 | 8 |
|  | 35-44 | 8 |
|  | 45-54 | 4 |
|  | 55-64 | 5 |
|  | ≥65 | 1 |
|  | Information not available | 1 |
| Sex | Female | 24 |
|  | Male | 6 |
| Sexuality | Heterosexual | 12 |
|  | Bisexual / Gay / Lesbian | 6 |
|  | Information not available | 12 |
| Ethnicity | Black British /Caribbean | 2 |
|  | British Asian / Indian | 3 |
|  | Chinese | 4 |
|  | Mixed Race | 2 |
|  | Romany | 1 |
|  | White British / English / Other | 18 |
|  | Information not available | 0 |
| Region of England (ONS classifications) | North West | 2 |
|  | North East | 1 |
|  | Yorkshire / Humber | 2 |
|  | West Midlands | 2 |
|  | East Midlands | 3 |
|  | South West | 2 |
|  | London | 17 |
|  | South East outside London | 1 |
|  | Information not available | 0 |

\* Diagnoses included: anxious avoidant personality disorder, borderline personality disorder, emotionally unstable personality disorder, narcissistic personality disorder, obsessive compulsive personality disorder, paranoid personality disorder and schizotypal personality disorder. Some participants also received earlier/concurrent diagnoses such as adjustment disorder, autism, bipolar disorder, depression, anxiety, eating
disorder, or been mis-diagnosed with bipolar, depression or schizophrenia. Some had their type of 'personality disorder' diagnosis changed or replaced by complex post-traumatic stress disorder.

### Findings

Results are reported in line with the thematic conceptual map (Fig 1), which presents the themes encapsulating how community services can best support people with CEN. The central theme in the map – Relational Practice – ties together all the other themes which describe how the approaches of individual staff as well as organisational structures and practices can operate in a way that supports positive interpersonal connections in the therapeutic relationship and provides a collaborative framework for the delivery of consistent, holistic and personalised care.

**Figure 1:**
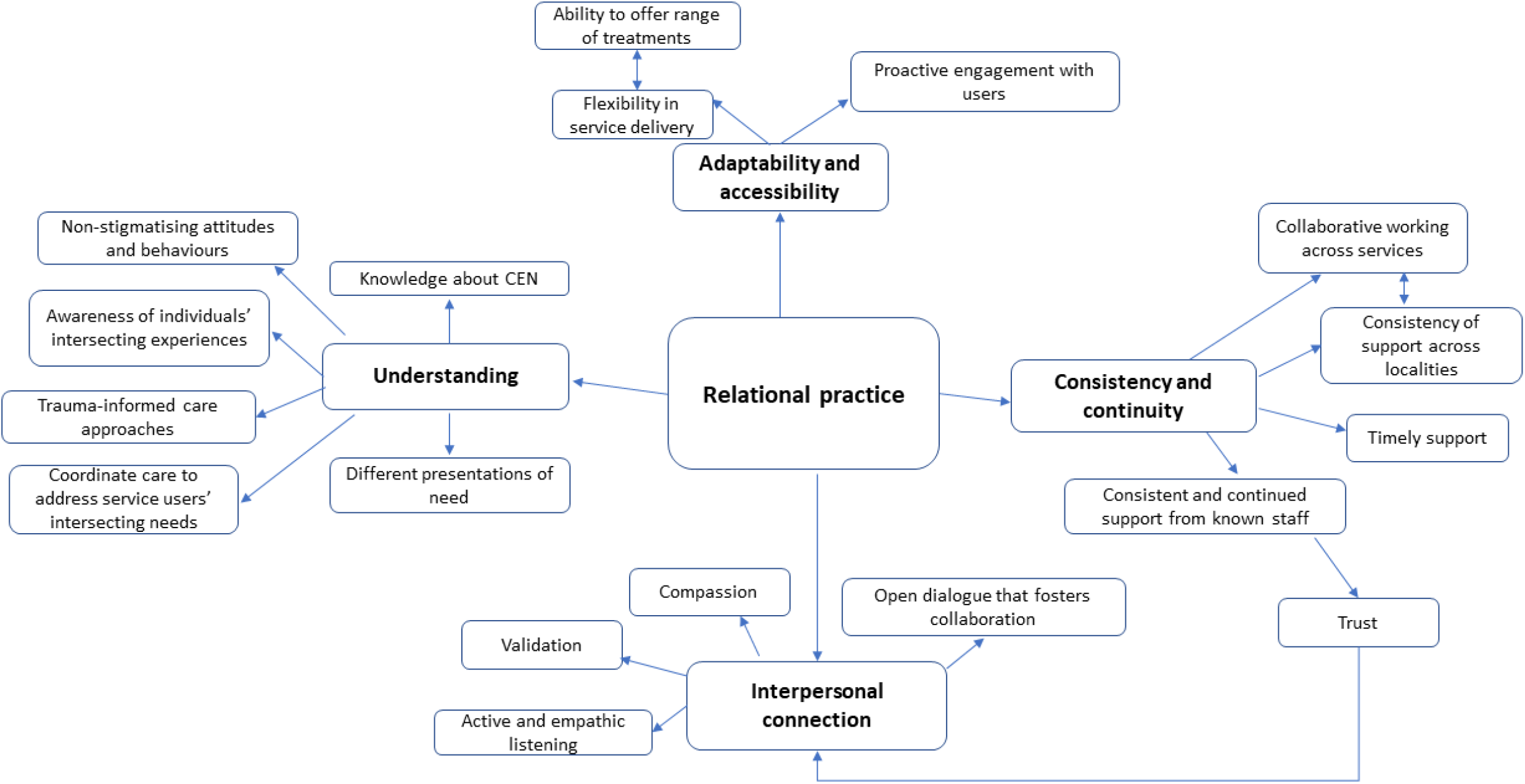
Conceptual Map.

### Understanding

The theme *Understanding* describes the need for community services to improve staff knowledge about CEN and to ensure staff adopt non-stigmatising and non-judgemental approaches. It describes the importance of services recognising service users’ intersecting identities (e.g. ethnicity, class, sexuality, disability) and how these may lead to additional disadvantage. It explores the need for services to address the multiple needs of service users (e.g. social, economic, housing) and the value of services adopting a trauma-informed approach to care, which acknowledges trauma experiences and which adopts practices to promote peoples’ safety, empowerment and choice.

#### Staff knowledge about CEN

Participants wanted to be supported by staff who were knowledgeable about their CEN and how to effectively support people with these needs:

> *“Having…the right workforce I think is really important. Even before you get to what services should be in place and where should they be, there is a how they should be operated, and that should be from…well informed practitioners”*

Yet many participants described experiences of being seen by staff in non-specialist community mental health services who were not knowledgeable about how to support someone with CEN:

> *“It’s not a well understood condition, either by the general public or by medical professionals. I think that’s obviously a huge weakness, that people just don’t know about or understand it”*

#### Non-stigmatising attitudes and behaviours

Several participants reported stigmatising attitudes among some staff, with respect to the diagnostic label ‘personality disorder’ and how it is often perceived, including being seen as someone who cannot be helped, being de-personalised or being viewed as a potential trouble-maker. These experiences could be pathologising and harmful. Examples were seen across generic mental health and voluntary sector services:

> *“People still have this attitude that basically there’s a group of people who are just impossible to work with and who will sabotage whatever you do”*
>
> *“There is a disparity in services, but I think that is due to either the stigma placed on people as they enter the door, or whom they come across and how they are perceived…I think there is that, sort of, dismissal of who you are, and not seeing the person as a person. They just see the diagnosis”*

Linked to this, participants commented that some staff view the needs of people with CEN as too challenging and so adopt dismissive or rejecting attitudes:

> *“It was really quite detrimental and actually harming when, rather than just saying, “We’re finding this hard to deal with*.*”…It felt like I was being blamed for the fact that my needs couldn’t be met”*

These perceptions resulted in some participants being denied access or turned away from a range of mental and physical health services:

> *“My experience of services is largely one of being dismissed or discriminated against on the basis of my diagnosis. I’ve had that from all kinds of people, from dieticians, to psychiatrists, to psychotherapists”*

The impact of these negative experiences left many participants feeling unheard and for a few it led them to disengage with services, as they felt it was causing them emotional harm:

> *“I didn’t feel my voice was being heard. I actually broke down contact with them because I thought they were making me worse. I just thought I could live it out by myself”*

Distinctions in how services responded to participants were made between generic mental health and specialist ‘personality disorder’ services:

> *“Because they are specialist services you get less of the stigma, I think, than you do in the general CMHT…they [specialist services] know…the things that you are likely to struggle with, but you still feel like an individual…rather than…”You are this kind of person*.*”*

#### Awareness of individuals’ intersecting experiences

Participants spoke of the importance of staff educating themselves about the intersecting identities that service users may have and how these may impact on an individual’s mental health:

> *“I am not expecting my therapist to have the same sexuality as me, but just being aware of the barriers, or the bi-phobia that a bi person can face, being aware of the different phobias”*
>
> *“Having cultural intelligence and cultural awareness…so I am not having to explain why when somebody said something did it hurt me…I need to acknowledge how it hurt me, and sit with that, versus feeling like I’m having to convince somebody that my pain is justified”*

Several participants reported experiences of discrimination connected to intersecting identities such as ethnicity, sexuality, age, class, physical appearance, and mental health need:

> *“It’s to do with your ethnicity, you know, even your body size, your age, all sorts make you even less likely to be listened to. That’s why I don’t like telling my age and I don’t like it when people ask about my ethnicity because they use all these things against you*.*”*
>
> *“Maybe it’s to do with being Black and just not being seen as trustworthy or being seen as, if I’m getting upset, as being aggressive when context isn’t taken into account. I feel like a lot of the stuff that’s happened wouldn’t have happened if I hadn’t been Black”*
>
> *Describing the text of a letter written by a community service* *“[It was] totally divorced from the context…”So, “[participant] is very angry and volatile*.*” Not, “[participant] is very angry and volatile because he lives month by month and will he be able to cover the rent?*…*and he’s really trying his hardest to get back to some so-called normality. That’s why he’s angry and volatile because he’s getting no help and support from anyone”*

Some participants spoke of needing to educate services about their intersecting identities and how they adversely impacted on their mental health:

> *“A huge part of my therapy became about me educating my therapist about a lot of the different things I had faced, or why it traumatised me, or why it affected me, because culturally we were different; ethnically we were different; different backgrounds, working class”*

#### Trauma-informed care approaches

Participants described the benefit of receiving trauma-informed care and called for wider adoption of this approach across community services. They described trauma-informed care as an understanding among staff that many people have developed CEN in response to past traumatic experiences and that changes in their behaviours often represent emotional responses to trauma. It provides a shift away from asking people “what’s wrong with you?” to a more person-centred, validating and sympathetic approach of “what’s happened to you?” It is an approach that demonstrates compassion and also seeks to ensure people feel safe (e.g. establishing clear communication channels with service users, being aware of the physical space of meeting rooms and whether they are comfortable, inviting and not overly clinical, asking people where they’d like to sit in a meeting room):

> *“After the first few sessions [trauma informed therapy], I felt like an explosion of knowledge in my learning and understanding*..*It’s very amazing”*
>
> *“trauma informed care…it acknowledges that…many people that suffer from personality disorder have done so because of the presence of early or indeed subsequent trauma in their life…This is a far more sympathetic and less judgemental way to proceed, and I look forward to when it is more mainstream than it currently is at the moment”*

Several participants described how their experiences of trauma were unacknowledged or even dismissed by services and described how organisational practices could inadvertently mirror abusive experiences:

> *“I was told very clearly that my history of trauma wasn’t relevant”*
>
> *“We’ve had so much instability in our lives [people with complex emotional needs]… Then when you go into services and services are chaotic or unstable, they [service users] don’t trust it”*

Another participant described how a lack of staff awareness of service users’ intersecting identities and non-adoption of trauma-informed care may result in misdiagnosis for some people:

> *“A few nurses have said to me there are some Black men that…go around, keep using services…It’s clear that there is some trauma going on. But they’ve never had the diagnosis. Because they will get something else as the diagnosis or…seen as the perpetrator, not a victim”*

#### Coordinate care to address service users’ multiple needs

Participants want services to recognise the impact of their individual, interconnecting needs on their mental health, including not only needs related to symptoms but also to social needs and wider problems in living:

> *“People [staff] will look at things like medication and therapy but life is much more than those two things. You know, how lonely people are…I think [services] needs to look at all elements of your life”*
>
> *“I think the priority is definitely the practical stuff…when I was homeless that was the priority…I really wasn’t in a place to be dealing with the kind of psychological stuff”*
>
> *“quite a lot of people have got financial problems, quite a lot of people have got housing problems. All these things are massively linked to mental health”*

In many cases, service providers did not appear to consider these factors, and this meant the type of support offered to participants was insufficient:

> *“The trauma of trying to keep the roof over my head and literally staying alive and eating and trying my best to stay off drugs…Every day was like a battle for mental and physical survival. It’s like none of those things actually matter…They never seem to be able to take these things into context of how this might affect you in your mental health”*

Some participants described positive experiences of staff across generic, specialist and voluntary sector services working proactively to understand and address the full range of their needs:

> *“She [care-coordinator] probably did more than any therapist I’ve had to turn me around… bringing to my attention educational courses, that she felt would assist me…*.*then able to intervene with the Local Authority…to change some of the appalling living conditions that I was surviving in at the time”*
>
> *“The [voluntary sector service programme] did offer…support with practical stuff which was, like, around social activity and social prescribing…how to do stuff…[it] did actually help”*

#### Different presentations of mental health need

Participants described how some services failed to identify and acknowledge the extent of their mental health need because of their physical and verbal presentations; seen as suggesting that they were better able to cope than was the case:

> *“The fact that I’m articulate and that I make eye-contact, that I dress and I wash, has been the biggest barrier to me to getting care…I said, “Look, I may not have gone out of the house for five days but because I have an appointment, I’ve done what most people would do. But that doesn’t mean that I’m well*.*”…I knew they didn’t get it”*

Consequently, several participants needed to advocate for themselves when engaging with services, but this approach often reinforced the perception that they were better able to manage their distress than was the case:

> *“There is this mismatch, I think, between me and my presentation, and me saying, “This is my distress. I need some help with this…because I am able to do that the expectations are really high…that I should be able to resolve that and then it leads to this tension”*

### Interpersonal connection

The theme Interpersonal Connection describes the need for staff to work compassionately and listen with empathy, to validate service users’ experiences and create an open dialogue with them.

#### Validation and active/empathic listening

Participants described how valuable and transformative it was when they interacted with staff who demonstrated active and empathic listening and who validated their experiences. These responses made them feel heard, cared for and supported. Such responses were reported across a range of non-specialist and specialist services:

> *“Accessing the mental health service and them actually saying, “I hear you. I hear that you need help*.*” That was, yes, it was very transformative”*
>
> *“Things I found helpful were, yes, people [voluntary sector service] listening, people empathising, people sympathising…People checking in, also, with what you need”*

#### Compassion

Participants wanted to receive support from staff who showed a genuine interest in understanding their distress and in identifying ways to best support them as an individual:

> *“Having a sense that someone actually really values you and is bothered about you, rather than that they’re trying to manage you in some way that they’ve been told is the right thing to do. So someone actually being responsive on an individual level”*

#### Open dialogue that fosters collaboration

Participants spoke of the benefit of an open dialogue with staff and collaborative discussions about their experiences/needs so that the right support is offered:

> *“Instead of making an assumption based on the notes that proceeded me…she said, “I’m not going to read anything. Let me get to know you first”… what that enabled was more of a dialogue… It facilitated a change in diagnosis. It facilitated a change in direction in terms of intervention…It’s such a simple thing, but it was transformative in my life”*

Many, however, reported staff not listening:

> *“It feels like I talk, and I’m not listened to, and then I’m just told, “This is what we are going to do*.*”*
>
> *“I was ignored…what she [staff at CMHT] was offering was, like, some breathing techniques. This is probably helpful for someone with lower and less complex needs than mine”*

### Consistency and Continuity

The theme Consistency and Continuity acknowledges the long-term mental health needs that people with CEN may have and how they would benefit from consistent, ongoing support from staff they know and with whom they have a working relationship. This construct also explores fragmentation of services across the statutory mental health care pathway and differences in service provision between localities.

#### Consistent and continued support from known staff

Participants wanted services to recognise the longer-term needs of people with CEN and to address these needs by providing consistency and continuity of care. They spoke of the value of services understanding that they have ongoing needs that require support not solely during periods of crisis:

> *“That stability, that consistency of care, and that understanding and approach that actually this is a long-term issue…I can operate with periods of health, but that doesn’t mean that it isn’t really hard and that I don’t need that support. As opposed to, “Oh, you know, you are doing really well at the moment. Off you pop*.*”*

Yet, several participants felt they were only provided with adequate periods of care during times of mental health crisis:

> *“Why should I have to be constantly on a, sort of, cliff edge, or jumping off the cliff for it to be resolved, or to have some intervention?”*
>
> *“I feel myself going up and down sometimes, so it’s frustrating that that support is not maintained…I am still suffering”*

Some participants described receiving consistent support from a member of staff that they had time to develop a relationship with, and who continued to support them over a longer period. This experience fostered trust and led participants to feel that services understood the longer-term nature of their needs and were invested in helping them address these:

> *“The thing that has been helpful is that in healthier periods whilst I haven’t needed the same intensity of care, having that continuity of care has kept me well, as opposed to then withdrawing and me deteriorating and then needing something more intense…definitely that stability of care, and that ability to be alongside you”*

Some participants described the shift patterns characteristic of crisis services as incompatible with providing consistency and continuity of care:

> *“If you’ve got a mental health team coming to visit you, you see a different person every day…you don’t get a chance to build a relationship with someone…if you’ve got a personality disorder or complex needs, that’s going to be more important than ever, to have one person to build a relationship with. Otherwise, everything is changing and that’s not what you need”*

#### Collaborative working across services

Participants described the benefit of agencies who provide mental health care working more collaboratively together to coordinate service users’ needs:

> *“The main improvement would be to join things up a bit better. Some of what you need is out there, it’s just that it’s not connected…the Crisis Resolution Team isn’t connected to my GP or to the [specialist unit in London]”*
>
> *“You’ve got to work together, because ultimately, by working together, you get that person well faster”*

Many participants spoke about their experience of statutory mental health services being disjointed, with discrete interventions delivered by each service and a lack of joined-up collaborative working practices:

> *“I kept getting bounced backwards and forwards between different bits of this, like, very opaque system…*.*it was a long process of being passed between different teams, and then, eventually, someone…saying “You should be treated in a different setting”…she referred me to [specialist community ‘personality disorder’ service]”*

#### Consistency of support across localities

Participants spoke of geographical disparities in the provision of mental health services, service availability and different approaches to care:

> *“In the south, you know, [I’ve] actually seen a crisis house … there’s none of that in this area … the language they use, the way the operate, it’s totally different depending on which part of the country you live in”*

#### Timely support

Participants spoke of the importance of being able to receive the right care at the right time, but found that high thresholds for access to some services made this difficult:

> *“To tell somebody in emotional pain, “Your pain is not significant for me compared to this person*.*”…Why do I have to be so far gone to get help?*

Some participants described how the tiered system required them to get a referral from one service to access another that provided more intensive mental health support. This process created delays, as service users needed to get approval for the referral from the first service and then wait for the documentation to be processed and approved by the second:

> *“The whole referral process [to a specialist community ‘personality disorder’ service] took six months…I was very, very mentally unwell…but unfortunately it felt like the policies and procedures were above everything else…it was very frustrating”*

### Adaptability and Accessibility

The theme Adaptability and Accessibility describes the value of services proactively engaging service users, implementing formalised peer support roles and supporting them to access a range of treatments. This theme also underlines the benefit of services adopting a flexible approach in their care of service users to ensure they receive the right support.

#### Proactive engagement with service users

Participants reflected on the benefit of services taking measures to promote engagement, through proactive steps to connect with service users:

> *“With this complex needs service, there is recognition that there may be many reasons why people aren’t turning up…They will say, “Why didn’t you come? What were the problems? How can I help you overcome those?”*

Some participants spoke of cathartic and healing experiences of receiving support from peer workers and called for peer-support roles to become formalised roles of employment and for peer-workers to receive skills-based training:

> *“I think the best people are people that have lived experience of mental health. Because…they can actually say, “I understand”, or, “I’ve been there,”…and they mean it, it’s true”*
>
> *“I feel very strongly that there needs to be…professionalisation of peer support…It is actually about really valuing the experiences of somebody with lived experience…it needs to be done in a framework of recompense for the work and appreciation of the role…I think it is really important, but I think it needs to be valued appropriately and invested in appropriately”*

#### Flexibility in service delivery

Participants spoke of the benefit of services being flexible in their approach to care to ensure that service users receive the right level of support at the right time. This included flexibility with respect to the types of support/treatment offered, with services having the ability to connect service users to different forms of support when needed, and in the ability of services to provide greater input at times when service users demonstrate greater need for support. These responses led to improvements in their well-being:

> *“[Service was] flexible around if I wasn’t getting on with a worker or if something just wasn’t right. I was getting CBT therapy for a while that wasn’t particularly right, and they referred me onto a psychodynamic psychological therapy. So, they were really good at being flexible”*
>
> *“She said [A&E psychiatric liaison], “Look, I’m going to give up my time, and you are going to come and see me here, at this hospital …I want to help you…let’s make something happen.”…She ended up seeing me…10 weeks in total, that was the beginning not just [of] me not trying to kill myself, but…the complete turnaround of my life was kickstarted by the positivity of just someone making that effort on my part”*

#### Ability to offer a range of treatments

Participants wanted services to facilitate their access to a range of forms of support. However, many explained that non-specialist services had limited knowledge of the support available for people with CEN or an understanding of the types of support that would be most appropriate for them:

> “*The whole system needs to be much more connected and people [say], “Okay, well, I know what I can help with and I know what I can’t help with*.*”…so everybody in the system is like, “Okay, this isn’t technically what I can help you with, but this person can*.*”*
>
> *“I’m becoming aware that there’re quite a lot of different tools out there that might work differently for different people at different times. Having the facility to actually, maybe, play around with those and experiment…rather than, “This is all that’s on offer, get on with it*.*”*

Participants recognised that the ability of services to deliver the good care practices outlined in the themes Consistency *and Continuity and Adaptability and Accessibility* were impacted by resourcing issues, service funding, staffing levels and staff training.

## Discussion

Participants reported a range of experiences of community practice from ‘abusive, damaging’ to ‘phenomenal’. Although participants reported several examples of positive care, many reported a range of negative experiences related to severe stigmatisation, a lack of staff knowledge and understanding of their intersecting needs/experiences, limited effective support and service fragmentation. *Relational Practice* was identified as the best way to support service users in the community, and participants reported elements of this practice across both specialist and non-specialist community services. This overarching theme includes four sub-themes: (1) Understanding; (2) Interpersonal Connection; (3) Consistency and Continuity; (4) Adaptability and Accessibility. Relational practice describes how staff and service practices/structures can work in a way that creates positive experiences for service users and leads to improved service user outcomes. It comprises staff delivering care in a non-stigmatising, individualised, compassionate, and trauma-informed manner. When staff work holistically and collaboratively with service users to coordinate support for their complex needs. When service structures allow for flexibility and continuity of care, accommodate the ongoing and changing nature of service users’ needs, and implement joint-working practices with other services. To successfully deliver relational practice, service design and intervention development must be co-produced with people with relevant lived experience, their carers and the professionals who support them.

### Understanding

#### Inclusive, non-judgemental, and non-discriminatory approaches

A key finding of this study is the need for services to adopt inclusive, non-judgemental and non-discriminatory approaches when supporting people with CEN. Participants reported numerous experiences of stigmatising attitudes from service providers regarding the diagnostic label ‘personality disorder’ and how it is often perceived, including being seen as someone who cannot be helped, being de-personalised or being viewed as a potential trouble-maker. There is evidence to suggest that people with CEN experience more stigma than people with other mental health diagnoses (17). Stigmatisation can result in iatrogenic harm when service providers dismiss peoples experience of distress, refuse them access to services or fail to demonstrate compassion in the therapeutic relationship (18). Generally, participants in this study who received support from specialist community ‘personality disorder’ services felt less stigmatised and judged about their mental health needs. This may reflect a better trained workforce with regards to knowledge and understanding of CEN (6).

Despite a body of literature evidencing the extent and impact of stigmatisation on people with CEN, few wide scale coordinated efforts have been undertaken to challenge these attitudes and practices. Our related qualitative work with community staff highlights that stigmatising views and behaviours arise not simply due to a lack of knowledge but also due to factors such as staff workloads, confidence and competencies in managing the multiple and longer-term needs of service users, as well as in managing issues of risk (Foye et al, submitted). There is an urgent need for research to identify the origins of these stigmatising behaviors and evaluations of programmes that seek to challenge and change stigmatising views and behaviours among staff. The Knowledge and Understanding Framework programme is an example of a training programme that has been shown to improve staff competencies in supporting service users with CEN (19).

Participants in this study also described how their intersecting identities (e.g. ethnicity, sexuality, age, class, physical appearance) can influence their mental health wellbeing and lead them to experience multiple forms of stigmatisation. Yet, participants reported that community services often demonstrated a lack of awareness of these issues or indeed perpetuated these forms of stigma. Across a range of healthcare settings, people are found to experience intersecting forms of stigma related to their different identities, which negatively impact on their physical and mental health (20). However, there remains a lack of research on the impact intersecting stigma experiences among people with mental health problems (21). There is some research describing how experiences of racism adversely impact on the ability of services to foster communication and trust, and in delivering effective care for Black and minority ethnic service users (22–24) which can lead to minority groups disengaging from mental health service use (25). There is also some research which shows that LGBTQ+ groups can experience a lack of understanding from staff when accessing healthcare (26), which our study provides further evidence of.

#### Holistic approaches

Our study adds further evidence to support our qualitative meta-synthesis finding that a holistic approach to care is of central importance to service users, and an overriding focus on the provision of psychological treatments targeting self-harm can neglect many current challenges that service users are dealing with (6). Participants in this study described wanting help with a range of social needs (e.g. housing, benefits, employment, social connections) alongside their mental health needs. They explained that a failure of services to recognise and address their competing social needs often meant they were offered forms of support that were either insufficient or with which they could not engage. Our review paper on models of care for people with CEN and the NICE guidelines for people with ‘borderline personality disorder’ also underline the need for treatment models to extend beyond just psychological therapies, to incorporate support for social and practical needs (14). Despite a well-established relationship between social and mental health, existing research and service guidelines predominately focus on the provision of psychological and pharmacological interventions (27). Further research would, therefore, benefit from reviewing the international evidence-base on integrated mental health and social need treatments and their respective strengths and weaknesses in improving service user experiences and outcomes.

#### Trauma-informed care approaches

Participants wanted community services to acknowledge their experiences of trauma. There are increasing calls by people with relevant lived experience, clinicians, research and policy makers for mental health services to adopt trauma-informed care (TIC) practices (10, 28–30). For example, the Power Threat Meaning Framework published by the British Psychological Society seeks to understand mental distress from a social, cultural, psychological and biological approach, replacing medicalised questions like “what is wrong with you?” to “what has happened to you?” It also acknowledges the centrality of the relational context in decisions about mental health need (31). Successful TIC approaches require responses at both organisational and individual staff levels (29). Organisational activities such as engaging service users in service design, hiring trauma-informed staff and training all staff in TIC practice, alongside staff implementation of routine enquiry about trauma, working collaboratively with service users in care planning and knowledge of trauma-based support services are critical (32). Further research is needed to identify how best to implement TIC approaches in mental health care settings.

### Interpersonal Connection

We found that service users want to be supported by staff who treat them with respect and compassion, who listen with empathy and who validate their individual experiences. These findings are supported by the wider qualitative literature on service users with CEN (6), and staff themselves also report the importance of these values and characteristics in their care of service users (7). These experiences can foster trust between service users and staff, critical for establishing and maintaining the therapeutic relationship (33).

Participants also want open dialogue and collaboration with staff to facilitate fuller disclosure of their experiences and needs and to ensure appropriate care is provided. Recent psychiatric guidelines on best practice approaches for people with CEN advocate for care management plans that are co-constructed by staff and service users, to ensure they fully reflect and address service users’ biological, psychological and social needs (10). Shared decision making is another approach that allows service users and staff to make joint healthcare decisions, and is found to improve service user outcomes (34). A recent review of shared decision making found that medical and diagnostic models of working in mental health services can result in staff not acknowledging service user expertise in their own mental health needs, and create power imbalances with service users feeling they are informed rather than involved in the care they receive (35).

### Consistency and Continuity

Participants wanted community services to acknowledge the long-term nature of CEN and to provide service users with consistent, ongoing support from staff. The provision of intensive psychological therapies (e.g. dialectical behavioural therapy) was beneficial to several participants but this treatment alone was not sufficient in addressing all of participants’ changing and longer-term needs. Participants identified that the delivery of longer-term support and consistency of service provision across different regions is often compromised by a lack of sufficient funding for services. This reflection aligns with the experience of clinicians, who report that a focus on avoiding dependency in the care relationship has encouraged the delivery of discrete interventions designed to promote rapid movement through and out of the mental health system, which is unsuitable for service users with enduring needs (36). Our related qualitative review exploring staff experiences in supporting people with CEN has identified a concern among some staff that offering sustained support can make the ending of treatment more challenging for service users (7). Yet, participants in this study reported that it is the absence of longer-term support which is particularly challenging. Further research would benefit from working with service users and staff to jointly identify best practice continuity of care approaches.

### Adaptability and Accessibility

Participants want services to be flexible with respect to the extent and type of support they offer service users, in response to their changing needs at any given time. This finding and those outlined above underline the benefit of flexible services structures which allow people to transition between more intensive and less intensive periods of support in a timely manner whilst also continuing to receive assistance for their ongoing needs. Participants also want staff to facilitate their engagement with services. Participants reflected that the ability of services to deliver these types of care is often compromised by a lack of sufficient funding, resulting in limited treatment options and limited staff capacity to work flexibly and proactively engage service users. Increased equity in service provision for people with CEN, compared to people with other mental health needs, would go some way to redress these limitations. Several participants also spoke of the value in services establishing peer support roles. Peer support approaches are increasing within mental health care settings but there is an absence of high-quality evaluations on the effectiveness of these models in improving service user outcomes (37) and warnings from some commentators that the formalisation of peer roles may threaten the equality and independence of this approach which can be a key to its success (38). Alongside the need to formally test peer support treatments of high methodological quality, further research could explore how peer support can best fit within services that model the positive care approaches outlined above (e.g. individualised, holistic and compassionate care that includes collaborative care planning and consistent and flexible support over time).

### Relational Practice

There are increasing calls for mental health services to adopt relational practice approaches, which prioritise the development and maintenance of therapeutic relationships over standardised procedures (36). Yet, it is argued that relational approaches may conflict with dominant managerialism practices (39) common in healthcare settings, as the latter emphasises implementation of standardised procedures and measurement of success against restricted performance targets (40). As a result, the importance of relational work in improving outcomes and care experiences can be downgraded. This study highlights the importance of relational work in improving service users’ experience of care and their outcomes. Other research supports this finding, with interpersonal components identified as key elements of evidenced-based treatments for CEN (41).The ability of staff to work relationally is shaped by organisational factors such as staffing ratios, supportive peer/management relationships (42) and opportunities for reflective practice and clinical supervision (10). In addition, an understanding at an organisational-level that relational work requires skills and expertise and are not simply innate characteristics of individuals (43).

### Strengths and Limitations

A group of experts by experience and occupation co-produced this study. In addition, most research interviews and analyses were carried out by lived experience researchers. We were successful in interviewing people with a broad range of experiences of service use with respect to specialist and non-specialist services, current and previous service use, service use over many years versus minimal use, including those who intentionally disengaged from service use after negative experiences. We did our best to diversify the study sample and 12/30 participants had ethnic backgrounds which are in the minority in England. There is a lack of research in this field with minority ethnic communities and this paper provides some valuable first-hand accounts. We also achieved a good geographical spread of participants; the East of England is the only region from which we had no representation.

Despite our broad inclusion criteria, all participants had received some sort of ‘personality disorder’ diagnosis at some time and we did not interview anyone who felt that they had been denied access to services because their CEN went undetected. We cannot, therefore, give any insight regarding the experiences of people who may have relevant needs, but do not access services or receive a diagnosis. Only a few men took part in the study (6/30). We asked participants about a broad range of community services, including voluntary sector services, but their views were largely based on their experiences of the NHS. Our recruitment methodology means that we have not captured the experiences of people who do not use social media or relevant CEN networks.

### Conclusions

This study identifies how relational practice approaches have the potential to improve community care for service users with CEN. Further research and service development is now needed and must be co-produced with people with relevant lived experience, their carers and the professionals who support them, to examine how best to implement relational practice approaches across the mental health service system.

## Lived Experience Commentary

This qualitative study reports significant difficulties in accessing appropriate community services for CEN which cast doubt on what progress, if any, has actually been made in this area since publication of the 2003 seminal document, “Personality Disorder: No Longer a Diagnosis of Exclusion” (44). We note the disturbing irony that the commissioning of this research has happened at a time when the coercive Serenity Integrated Mentoring (SIM) programme (45), which threatens to criminalise people with CEN in crisis and withholds essential mental and physical health care (46), has been adopted so readily across mental health trusts in England without proper scrutiny or evidence base.

When interviewing for this study, we were particularly struck by the recurrent accounts of stigmatising attitudes amongst staff in community mental health services. Encountering stigma in this context can cause profound iatrogenic harm and tackling this pervasive and enduring stigma must be a key priority for future service improvement. The paper’s overarching theme and recommendation of relational practice – a collaborative framework for providing care – may contribute to a sound foundation for addressing this.

Such collaboration in individual care reflects the need for staff to embrace true co-production at all stages from design to delivery of any services for people with CEN. However, that will require a substantial culture change in many services where the reality of co-production initiatives often falls far short of its guiding principles (47). Likewise, embedding lived experience practitioners at all levels of seniority will be indispensable for meaningful change, but this is often still met with great resistance from staff who fear their status being challenged.

The findings also highlight the need to acknowledge and address intersecting challenges of trauma, inequalities and discrimination in a more holistic approach. We suggest that future research and policy work need to go further than the commission for this project allowed, listen to the experiential knowledge of service users in this study and the project’s January 2019 workshop (48), and abandon the pejorative construct of the disordered personality which fuels stigmatising attitudes. There is an urgent need to develop approaches focused around what has happened to the person in distress and supporting them with their natural reactions to trauma without judgement or prejudice.

## Data Availability

The data that support the findings of this study are available on request from the corresponding author (KT). The data are not publicly available due to the sensitive nature of the data.

## Declarations

## Abbreviations

(CEN): Complex emotional needs
(LGBTQ+): Lesbian, Gay, Bisexual, Transgender, Queer/Questioning people

## Ethics Approval and Consent to Participate

Ethical approval for the study was obtained from King College London’s PNM Research Ethics Subcommittee on 16/05/2019, reference HR-18/19-10795. Informed consent was obtained from all study participants.

## Consent for publication

Not Applicable

## Competing Interests

The authors declare that they have no competing interests.

## Funding

This paper presents independent research commissioned and funded by the National Institute for Health Research (NIHR) Policy Research Programme, conducted by the NIHR Policy Research Unit (PRU) in Mental Health. The views expressed are those of the authors and not necessarily those of the NIHR, the Department of Health and Social Care or its arm’s length bodies, or other government departments.

## Authors’ contributions

KT, RS, EB, SJe, TJ, DA, JR, JB, MJC, RH, SM, VN, AS, BLE, SJo, SO co-produced the study protocol and interview topic guide. EB, SJe, DA, JR, RS, KT, UF led the data analyses and all authors conducted interpretation of the data. KT, RS, JO wrote the manuscript and EB, SJe, TJ, DA, JR, JB, MJC, OD, RH, PM, SM, VN, UF, AS, BLE, SJo, SO critically reviewed and improved the manuscript. All authors have approved the manuscript.

## Acknowledgements

We thank all of the service users who participated in this study and shared their experiences with us. We wish to thank all members of our study working group for their continued guidance and support throughout this project.

## Notes

### Competing Interest Statement

The authors have declared no competing interest.

### Clinical Trial

n/a

### Author Declarations

Ethical approval for the study was obtained from Kings College London PNM Research Ethics Subcommittee on 16/05/2019, reference HR-18/19-10795.

### Summary of Updates

This paper now includes a lived experience commentary of the work, following the Conclusions section

## References

1. Winsper C, Bilgin A, Thompson A, Marwaha S, Chanen AM, Singh SP, et al. The prevalence of personality disorders in the community: a global systematic review and meta-analysis. Br J Psychiatry. 2019;216(2):69–78.

2. Evans S, Sethi F, Dale O, Stanton C, Sedgwick R, Doran M, et al. Personality disorder service provision: a review of the recent literature. Mental Health Review Journal. 2017.

3. Tyrer P, Reed GM, Crawford MJ. Classification, assessment, prevalence, and effect of personality disorder. The Lancet. 2015;385(9969):717–26.

4. Crawford MJ, Rushwaya T, Bajaj P, Tyrer P, Yang M. The prevalence of personality disorder among ethnic minorities: findings from a national household survey. Personality and Mental Health. 2012;6(3):175–82.

5. Hossain A, Malkov M, Lee T, Bhui K. Ethnic variation in personality disorder: evaluation of 6 years of hospital admissions. BJPsych bulletin. 2018;42(4):157–61.

6. Sheridan Rains L, Echave A, Rees J, Scott HR, Lever-Taylor B, Broeckelmann E, et al. Service user experiences of community services for Complex Emotional Needs: A qualitative thematic synthesis.medRxiv. 2020.

7. Troup J, Lever-Taylor B, Rains LS, Broeckelmann E, Russell J, Jeynes T, et al. Clinician perspectives on what constitutes good practice in community services for people with Complex Emotional Needs: A qualitative thematic meta-synthesis. medRxiv. 2020.

8. Chanen A, Sharp C, Hoffman P, for Prevention GA. Prevention and early intervention for borderline personality disorder: a novel public health priority. World Psychiatry. 2017;16(2):215.

9. Flynn S, Graney J, Nyathi T, Raphael J, Abraham S, Singh-Dernevik S, et al. Clinical characteristics and care pathways of patients with personality disorder who died by suicide. BJPsych open. 2020;6(2).

10. Royal College of Psychiatrists. Services for people diagnosable with personality disorder. London: Royal College of Psychiatrists; 2020.

11. Simonsen S, Bateman A, Bohus M, Dalewijk HJ, Doering S, Kaera A, et al. European guidelines for personality disorders: past, present and future. Borderline personality disorder and emotion dysregulation. 2019;6(1):1–10.

12. NHS England. The NHS Long Term Plan. London: NHS England; 2019.

13. Ng F, Townsend ML, Jewell M, Marceau EM, Grenyer BF. Priorities for service improvement in personality disorder in Australia: perspectives of consumers, carers and clinicians. Personality and Mental Health. 2020;14(4):350–60.

14. Storebø OJ, Stoffers-Winterling JM, Völlm BA, Kongerslev MT, Mattivi JT, Jørgensen MS, et al. Psychological therapies for people with borderline personality disorder. Cochrane Database of Systematic Reviews. 2020(5).

15. Booth A, Hannes K, Harden A, Noyes J, Harris J, Tong A. COREQ (consolidated criteria for reporting qualitative studies). Guidelines for reporting health research: a user’s manual. 2014:214–26.

16. NVivo 12 [Internet]. QSR International Pty Ltd 2018.

17. Rao H, Mahadevappa H, Pillay P, Sessay M, Abraham A, Luty J. A study of stigmatized attitudes towards people with mental health problems among health professionals. Journal of psychiatric and mental health nursing. 2009;16(3):279–84.

18. Sheehan L, Nieweglowski K, Corrigan P. The stigma of personality disorders. Current Psychiatry Reports. 2016;18(1):11.

19. Davies J, Sampson M, Beesley F, Smith D, Baldwin V. An evaluation of Knowledge and Understanding Framework personality disorder awareness training: Can a co-production model be effective in a local NHS mental health Trust? Personality and mental health. 2014;8(2):161–8.

20. Turan JM, Elafros MA, Logie CH, Banik S, Turan B, Crockett KB, et al. Challenges and opportunities in examining and addressing intersectional stigma and health. BMC medicine. 2019;17(1):1–15.

21. Oexle N, Corrigan PW. Understanding mental illness stigma toward persons with multiple stigmatized conditions: Implications of intersectionality theory. Psychiatric Services. 2018;69(5):587–9.

22. Holley LC, Tavassoli KY, Stromwall LK. Mental illness discrimination in mental health treatment programs: Intersections of race, ethnicity, and sexual orientation. Community mental health journal. 2016;52(3):311–22.

23. Delphin-Rittmon ME, Flanagan EH, Andres-Hyman R, Ortiz J, Amer MM, Davidson L. Racial-ethnic differences in access, diagnosis, and outcomes in public-sector inpatient mental health treatment. Psychological Services. 2015;12(2):158.

24. Gabbidon J, Farrelly S, Hatch SL, Henderson C, Williams P, Bhugra D, et al. Discrimination attributed to mental illness or race-ethnicity by users of community psychiatric services. Psychiatric Services. 2014;65(11):1360–6.

25. Richman LS, Kohn-Wood LP, Williams DR. The role of discrimination and racial identity for mental health service utilization. Journal of Social and Clinical Psychology. 2007;26(8):960–81.

26. Kattari SK, Walls NE, Whitfield DL, Langenderfer-Magruder L. Racial and ethnic differences in experiences of discrimination in accessing health services among transgender people in the United States. International Journal of Transgenderism. 2015;16(2):68–79.

27. Johnson S. Social interventions in mental health: a call to action. Springer; 2017.

28. Sweeney A, Clement S, Filson B, Kennedy A. Trauma-informed mental healthcare in the UK: what is it and how can we further its development? Mental Health Review Journal. 2016.

29. Substance Abuse and Mental Health Services Administration. SAMHSA’s concept of trauma and guidance for a trauma-informed approach. Rockville, MD: Substance Abuse and Mental Health Services Administration.; 2014.

30. Johnstone L, Boyle M, Cromby J, Dillon J, Harper D, Kinderman P, et al. The Power Threat Meaning Framework: Towards the identification of patterns in emotional distress, unusual experiences and troubled or troubling behaviour, as an alternative to functional psychiatric diagnosis.. Leicester: British Psychological Society; 2018.

31. Johnstone L, Boyle M, Cromby J, Dillon J, Harper D, Kinderman P, et al. The Power Threat Meaning Framework: Overview. Leicester: British Psychological Society; 2018.

32. Menschner C, Maul A. Key ingredients for successful trauma-informed care implementation: Center for Health Care Strategies, Incorporated Trenton; 2016.

33. Langley G, Klopper H. Trust as a foundation for the therapeutic intervention for patients with borderline personality disorder. Journal of psychiatric and mental health nursing. 2005;12(1):23–32.

34. Huang C, Plummer V, Lam L, Cross W. Perceptions of shared decision-making in severe mental illness: An integrative review. Journal of psychiatric and mental health nursing. 2020;27(2):103–27.

35. Castillo H, Ramon S. “Work with me”: service users’ perspectives on shared decision making in mental health. Mental Health Review Journal. 2017.

36. Dale O, Haigh R, Blazdell J, Sethi F. Social psychiatry, relational practice and learning from COVID-19. Mental Health Review Journal. 2020;25(4):297–300.

37. Lloyd-Evans B, Mayo-Wilson E, Harrison B, Istead H, Brown E, Pilling S, et al. A systematic review and meta-analysis of randomised controlled trials of peer support for people with severe mental illness. BMC psychiatry. 2014;14(1):1–12.

38. Faulkner A, Basset T. A long and honourable history. The Journal of Mental Health Training, Education and Practice. 2012.

39. Ingram R, Smith M. Relationship-based practice: emergent themes in social work literature. 2018.

40. Carlisle Y. Complexity dynamics: Managerialism and undesirable emergence in healthcare organizations. Journal of Medical Marketing. 2011;11(4):284–93.

41. Bateman AW, Gunderson J, Mulder R. Treatment of personality disorder. The Lancet. 2015;385(9969):735–43.

42. Doane GH, Varcoe C. Relational practice and nursing obligations. Advances in Nursing Science. 2007;30(3):192–205.

43. Carlson JH, Crawford M. Perceptions of relational practices in the workplace. Gender, Work & Organization. 2011;18(4):359–76.

44. National Institute for Mental Health in England. Personality Disorder: No Longer a diagnosis of exclusion, policy implementation guidance for the development of services for people with personality disorder. London: Department of Health; 2003.

45. Jennings P, Matheson-Monnet CB. Multi-agency mentoring pilot intervention for high intensity service users of emergency public services: The Isle of Wight Integrated Recovery Programme. Journal of Criminological Research, Policy and Practice. 2017.

46. StopSIM Coalition. Coalition Statements (StopSIM Coalition Consensus Statement relating to The High Intensity Network (HIN) and Serenity Integrating Mentoring (SIM)): StopSIM Coalition; 2021 [Available from: https://www.stopsim.co.uk/.

47. Slay J, Stephens L. Co-production in mental health: A literature review. London: new economics foundation. 2013:4.

48. National Institute for Health Research Mental Health Policy Research Unit. Community ‘Personality Disorder’ Services Research Workshop: National Institute for Health Research Mental Health Policy Research Unit; 2019 [Available from: https://www.ucl.ac.uk/psychiatry/research/nihr-mental-health-policy-research-unit/past-events/community-personality-disorder-services.

